# StartReact increases probability of muscle activity and distance in severe/moderate stroke survivors during two-dimensional reaching task

**DOI:** 10.1101/19012252

**Authors:** Marziye Rahimi, Claire F. Honeycutt

## Abstract

**Objective:** StartReact elicits faster, larger, and more appropriate muscle activation in stroke survivors but has been only cursorily studied to date during multi-jointed reaching. Our objective was to evaluate StartReact on unrestricted, two-dimensional point-to-point reaching tasks post-stroke.

**Method:** Data from 23 individuals with stroke was collected during point-to-point reaching. Voluntary and StartReact trials were compared between mild, severe/moderate, and the unimpaired arm.

**Results:** StartReact showed an increase in probability of muscle activity, larger muscle activity amplitude and faster muscle activity onset. Despite changes in muscle activity, metrics of movement (distance, final error, linear deviation) were largely the same between StartReact and Voluntary trials except in severe/moderate stroke who had larger reaching distances during StartReact.

**Conclusion:** While StartReact impacted many metrics of muscle activity, the most profound effect was on probability of muscle activity increasing 34% compared to Voluntary which allowed severe/moderate subjects to increase reaching distance but did not translate to decrease in final error suggesting that the additional movement was not always directed towards the appropriate target.

**Significance:** These results indicate that SR has the capacity to activate paralyzed muscle in severe/moderate patients, but future studies are needed to explore the possible use of SR in the rehabilitation.

## Introduction

The ability of the startle reflex to involuntarily release voluntarily planned movement, a phenomenon referred to as StartReact (SR), has been investigated extensively for the past 20 years (Valls-Solé et al. 1999). SR has been used as a tool to evaluate the underlying mechanisms of neurological diseases and injuries including stroke (Jankelowitz and Colebatch 2004; Honeycutt and Perreault 2012; Honeycutt et al. 2015; Marinovic et al. 2016; Choudhury et al. 2019), spinal cord injury (Jankelowitz and Colebatch 2004; Baker and Perez 2017), Parkinson’s (Carlsen et al. 2013; Nonnekes et al. 2014a), corticospinal degeneration (van Lith et al. 2018) and hereditary spastic paraplegia (Nonnekes et al. 2014b). SR has been shown to elicit faster, larger, and more appropriate muscle activation in individuals who have had a stroke (Honeycutt and Perreault 2012, 2014; Honeycutt et al. 2015; Marinovic et al. 2016). Specifically, when elbow flexion movements are elicited via SR, the onset latency and agonist/antagonist timing of stroke survivors are not statistically different from unimpaired subjects of the same age (Honeycutt and Perreault 2012). This result was replicated by an independent group (Marinovic et al. 2016) who additionally demonstrated that SR could be evoked using electrical stimulation in individuals with stroke. In addition to temporal characteristics, SR increased the amplitude of muscle activity, which is most striking in individuals with more severe impairment (Honeycutt and Perreault 2014). Similar results (decreased onset latency and increased muscle activity) are seen during SR in hand extension/flexion tasks post-stroke indicating that SR can influence movement across the entire limb post-stroke (Honeycutt et al. 2015).

The capacity of SR to alter muscle activation has led some to argue for the use of SR as a therapeutic intervention for individuals with stroke but SR’s capacity to change muscle activity during multi-joint, more functionally relevant reaching movement has only been cursorily studied to date. Most previous SR work has evaluated only single joint movement (Cressman et al. 2006; Maslovat et al. 2009, 2011; Carlsen et al. 2011, 2012; Alibiglou and MacKinnon 2012; Ravichandran et al. 2013; Castellote and Valls-Solé 2015; Sutter et al. 2016; Marinovic et al. 2017; Leow et al. 2018) including the previous work in individuals with stroke (Honeycutt and Perreault 2012, 2014; Marinovic et al. 2016). In unimpaired individuals, SR readily evokes multi-jointed reaching movements (Wright et al. 2015; Ossanna et al. 2019) and combined tasks such as simultaneous pinch and elbow flexion (Castellote and Kofler 2018) in an unrestricted workspace but individuals with stroke have abnormal muscle activity patterns and spasticity which casts doubt on if SR would be accessible during unrestricted reaching movements post-stroke and more relevantly generate quantitative changes that improve reaching performance in this population. Moreover, SR extension movements are interrupted by functionally inappropriate flexor activity disrupting movement to the intended target (Honeycutt and Perreault 2014; Tazoe and Perez 2017; Choudhury et al. 2019). This inappropriate flexor activity is more pronounced in individuals with severe impairment and has been shown to be largest in individuals with high spasticity. These abnormalities draw into question if SR would be effective, or even advisable, in severe/moderate stroke survivors.

Therefore, the objective of this study is to evaluate the impact of SR during unrestricted, two-dimensional point-to-point reaching tasks in individuals with a wide range of impairment – particularly severe and moderate stroke. We hypothesized that SR would decrease reaction times, increase muscle activation and muscle amplitude in all stroke survivors, and produce larger reaching distances in stroke survivors with severe/moderate impairment.

## Methods and Materials

### Subjects

Twenty-three individuals (age = 19 to 85 years) with chronic severe-to-mild stroke (UEFM = 8-66/66) and no-to-severe spasticity (Modified Ashworth = 0-4/4) participated in this study (Table 1). Inclusion/exclusion criteria were: no injury to arm/shoulder in past 6 months, at least 6 months post-stroke, no hearing loss/sensitivity, no dizzy or fainting spells, no seizure or heart attacks, measurable impairment in the upper extremity, and could not be pregnant. This study was approved by Arizona State University’s Institutional Review Board STUDY00002440. Subjects were informed of all potential risks prior to participation in the study and verbal/written consent was obtained.

**Table 1.** Summary of subjects’ characteristics

| Number | Sex | Age (years) | Duration from onset of stroke (years) | Paretic arm | UEFM Score | Modified Ashworth Score |  |  |  |
| --- | --- | --- | --- | --- | --- | --- | --- | --- | --- |
|  |  |  |  |  |  | Shoulder flexion | Shoulder extension | Elbow extension | Elbow flexion |
| 1 | M | 31 | 0.9 | R | 65 | 0 | 0 | 0 | 0 |
| 2 | M | 77 | 3.7 | R | 25 | 0 | 0 | 3 | 3 |
| 3 | M | 59 | 5.3 | L | 41 | 0 | 1 | 0 | 1+ |
| 4 | M | 36 | 10.4 | R>L | 31 | 0 | 0 | 3 | 2 |
| 5 | F | 66 | 4.7 | R | 66 | 0 | 0 | 0 | 0 |
| 6 | M | 61 | 1.7 | L | 55 | 1 | 1 | 1+ | 0 |
| 7 | F | 61 | 1.6 | L | 64 | 0 | 0 | 0 | 0 |
| 8 | M | 47 | 1.5 | L | 61 | 0 | 0 | 0 | 0 |
| 9 | F | 39 | 3.7 | R | 19 | 3 | 3 | 3 | 3 |
| 10 | M | 51 | 1.1 | L | 24 | 0 | 1 | 3 | 3 |
| 11 | M | 28 | 0.5 | R | 35 | 0 | 0 | 2 | 2 |
| 12 | M | 52 | 11.7 | L>R | 58 | 0 | 0 | 0 | 2 |
| 13 | M | 71 | 12.4 | L | 8 | 1 | 1 | 3 | 4 |
| 14 | F | 65 | 17.4 | R | 65 | 0 | 0 | 0 | 0 |
| 15 | F | 49 | 1.2 | L | 11 | 1 | 0 | 2 | 2 |
| 16 | F | 65 | 7.8 | L | 11 | 0 | 1 | 1+ | 0 |
| 17 | F | 19 | 0.8 | L>R | 14 | 0 | 0 | 0 | 1 |
| 18 | M | 70 | 2.5 | L | 64 | 0 | 0 | 0 | 0 |
| 19 | F | 33 | 13.5 | R | 11 | 1 | 1 | 1 | 1 |
| 20 | M | 68 | 3.3 | R | 53 | 0 | 0 | 0 | 0 |

### Protocol

Subjects were asked to sit comfortably in the experimental chair. Ag/Cl surface electrodes [MVAP Medical Supplies, Newbury Park, CA] were used to record activity from the brachioradialis (BR), biceps (BIC), triceps lateral head (TRI), pectoralis (PEC), anterior deltoid (AD), and posterior deltoid (PD) muscles. Electrodes were also placed on the left (LSCM) and right (RSCM) sternocleidomastoid muscles to monitor startle activity (Carlsen et al. 2003). EMG signals were amplified by the Bortec AMT-8 system [Bortec Biomedical, Calgary, Alberta, Canada]. This system had a bandwidth of 10-1000Hz, an input impedance of 10GΩ, and a common mode rejection ratio of 115 dB at 60Hz. Electromyography (EMG) data were recorded at gain of 1500 and frequency of 3000Hz by a 32-channel, 16-bit data acquisition system [NI USB-6363, National Instrumentation, Austin, TX].

For this study, the InMotion2 Interactive Therapy System (Interactive Motion Technologies, Inc, Watertown, MA 02472 USA) was used to record time and position data for the reaching tasks performed by the subject at a sampling frequency of 1000 Hz. The InMotion2 system is a commercial version of the MIT-Manus and is designed for use in a clinical environment (Hogan et al. 1994). The arm rested on a custom-made arm support attached to the robot arm. Both the impaired and unimpaired arms were evaluated for all subjects. Three subjects had a stroke in both arms. For these individuals, the most impaired arm was tested, and the least impaired arm was not included in analysis.

Subjects were instructed to perform reaching movements to three targets starting from an initial arm position of shoulder abduction at 70°, shoulder flexion at 40° and elbow angle at 90° (all ±5° to subjects comfort). A monitor in front of the subjects depicted location of the home and target positions as well as the location of their hand. The position of home circle was fixed. The distance between home and target circles was proportional to the subjects’ arm lengths. This distance was calculated using the formula

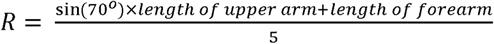 (Fig. 1).

Subjects were instructed to move following two soft (80 dB) auditory sounds. Subjects were told to plan to move after the first sound (GET READY) and reach as fast as possible after hearing the second sound (GO). GO cue was delivered between 2-3 seconds after the GET READY cue to prevent anticipation. The cursor disappeared right after they left the home circle and reappeared after 1 second. Subjects practiced reaching to each target 15 times before data collection. The cursor did not disappear during the practice time. There were two additional trial types: Classic startle and StartReact (SR) trials. During these trials the GET READY cue (classic startle) or the GO cue (SR) was replaced with a startling sound of 115dB. Subjects performed three blocks of arm reaches to each target. The order of each block was generated randomly. Each block contained 15 trials of the three different trial types: 1) Voluntary (VOL): In 66.7% of trials GO and GET READY were at 80 dB; 2) StartReact (SR) trials: In 33.4% of trials GO was at 115dB; 3) Classic startle trials: 3 trials per arm during the first block GET READY was at 115dB. Modified Ashworth Scales and Upper Extremity Fugl-Meyer assessments (UEFM) were collected at the end of the experiment.

### Data Analysis

EMG data were rectified and smoothed in MATLAB (R2017b) using a 10-point moving average. We calculated the following outcome measures: muscle onset, movement onset, movement distance, deviation from linearity, final error, mean muscle activity amplitude, and probability of muscle activity. Muscle onset latency was first detected using a custom MATLAB script that detected EMG activity greater than the background activity plus 3 standard deviations. Background was calculated using 500 ms prior to the GO cue. Visual inspection and corrections were conducted by an experimenter blinded to trial type. Movement onset was defined as the time when the subject left the 1 cm HOME circle. Movement distance was the distance travelled from the movement onset until the velocity of movement was found to be less than 0.0001 m/s (final position). We normalized the movement distance to *R* (Fig. 1). Deviation from linearity was defined using the following formula 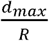 (Sainburg et al. 1993; Bagesteiro and Sainburg 2003; Schaefer et al. 2009). Final error was the distance between the final position and the center of the intended TARGET. Mean muscle activity amplitude was calculated as the mean EMG activity over the first 70 ms proceeding the onset of muscle activity. EMG amplitude was normalized to the percent of maximum voluntary muscle activity collected during the experiment. The probability of muscle activity for each muscle was calculated as the number of trials where an EMG onset could be identified divided by the total number of trials. Trials in which the subject failed to move or was distracted were eliminated from analysis (4.8 % of trials). Additionally, 3 subjects were eliminated because they did not exhibit at least 3 SR trials (measured as SCM activity prior to 120ms).

**Fig. 1.**
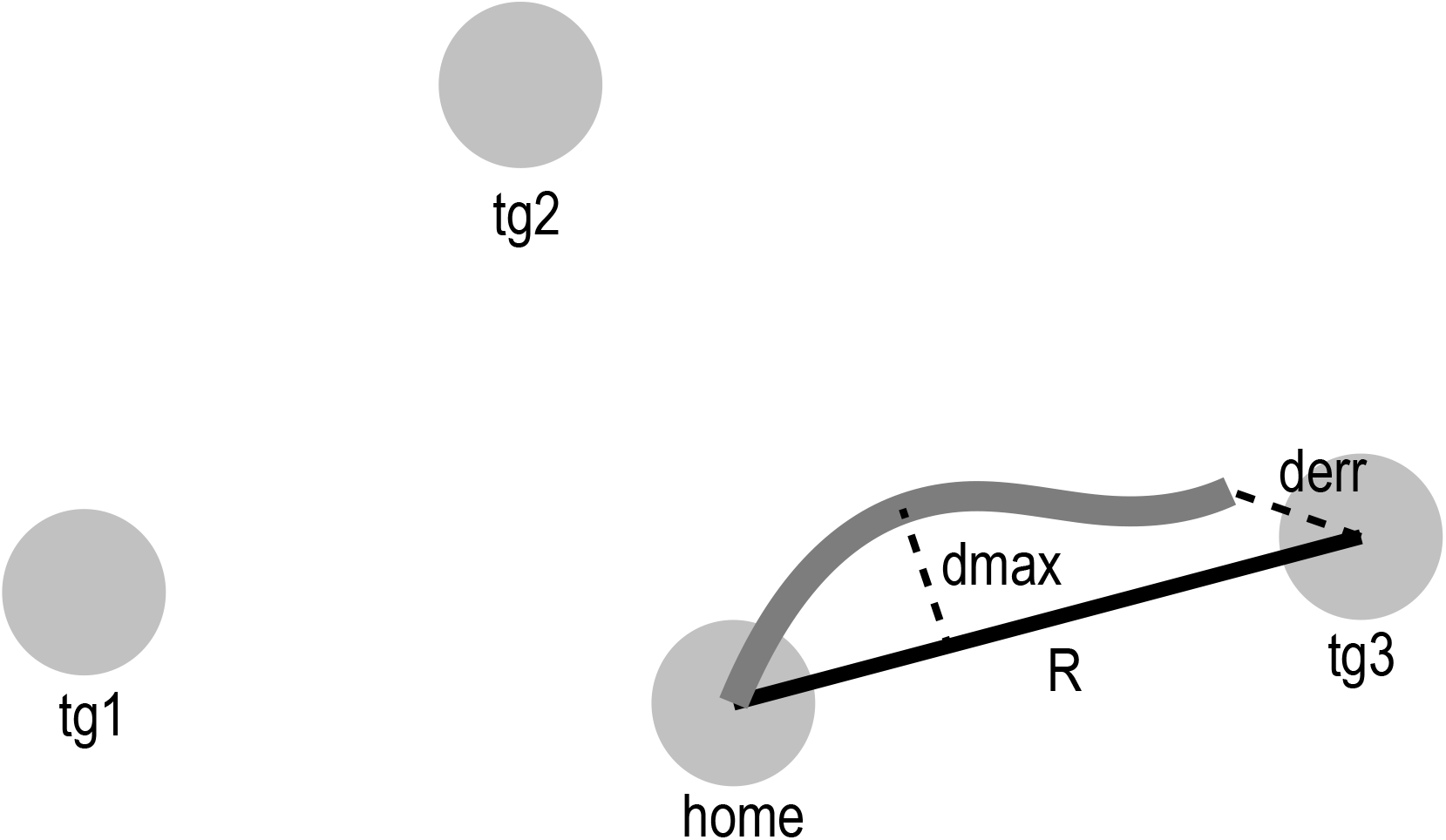
Target Positions. The locations of target and home circles for the right arm are presented. The left arm targets were the mirror image of these targets. *R* was the distance between the home circle and the target circles (black line). The gray line is the movement trajectory from the start point to the end point. d_max_ is the maximum distance between the movement trajectory and the axis connecting the home to the target. d_err_ is the distance between the target and the end point. tg: target.

### Statistical Analysis

Individuals with stroke were separated into two groups: 11 severe/moderate (UEFM < 45/66) and 9 mild (UEFM > 45/66). The unimpaired arm (of 17 subjects; 3 subjects had bilateral stroke) was evaluated for comparison. We used a Generalized Mixed Effects model (Pinheiro, JC.; Bates 2000) in R 2017 version 3.4.2 (Bates 2005) for all the comparisons. Dependent variables included all calculated metrics listed above (e.g. onset latency, movement onset, etc.). The fixed effects were condition (VOL, SR), population (unimpaired, mild, severe/moderate) and muscle (BR, BIC, TRI, PEC, AD, PD). Subject was treated as a random factor and 95% confidence was used to define statistical significance.

## Results

SR and VOL movements were similar in unimpaired and mild, but in severe subjects SR movements were larger and recruited more muscles (Fig. 2). Similar to previous work (Ossanna et al. 2019), SR and VOL movement trajectories to the target were not different (except onset latency) in the unimpaired and mild stroke groups (Fig. 2a and 2b). Typical of SR, muscle activity was faster with similar activation patterns for both unimpaired and mild. However, contrary to previous work in young adults (Carlsen et al. 2004; Ossanna et al. 2019), the SR amplitude of muscle activity was larger in unimpaired and mild stroke. In severe stroke, SR had a larger impact on movement trajectory compared to mild and unimpaired groups (Fig. 2c). During VOL, the representative severe subject #18 was unable to leave the HOME target and muscle activity was absent or significantly delayed. Conversely, SR movements left the HOME target and traveled away from the body. Similarly, muscle activity, absent during VOL, was present in all muscles at a fast latency during SR. It is important to note that traces seen in Fig. 2c represent SR movement trajectories to all 3 targets. Thus, while SR movements were larger and directed away from the body, subjects often did not achieve the target i.e. final error was not improved (see group results below).

**Fig. 2.**
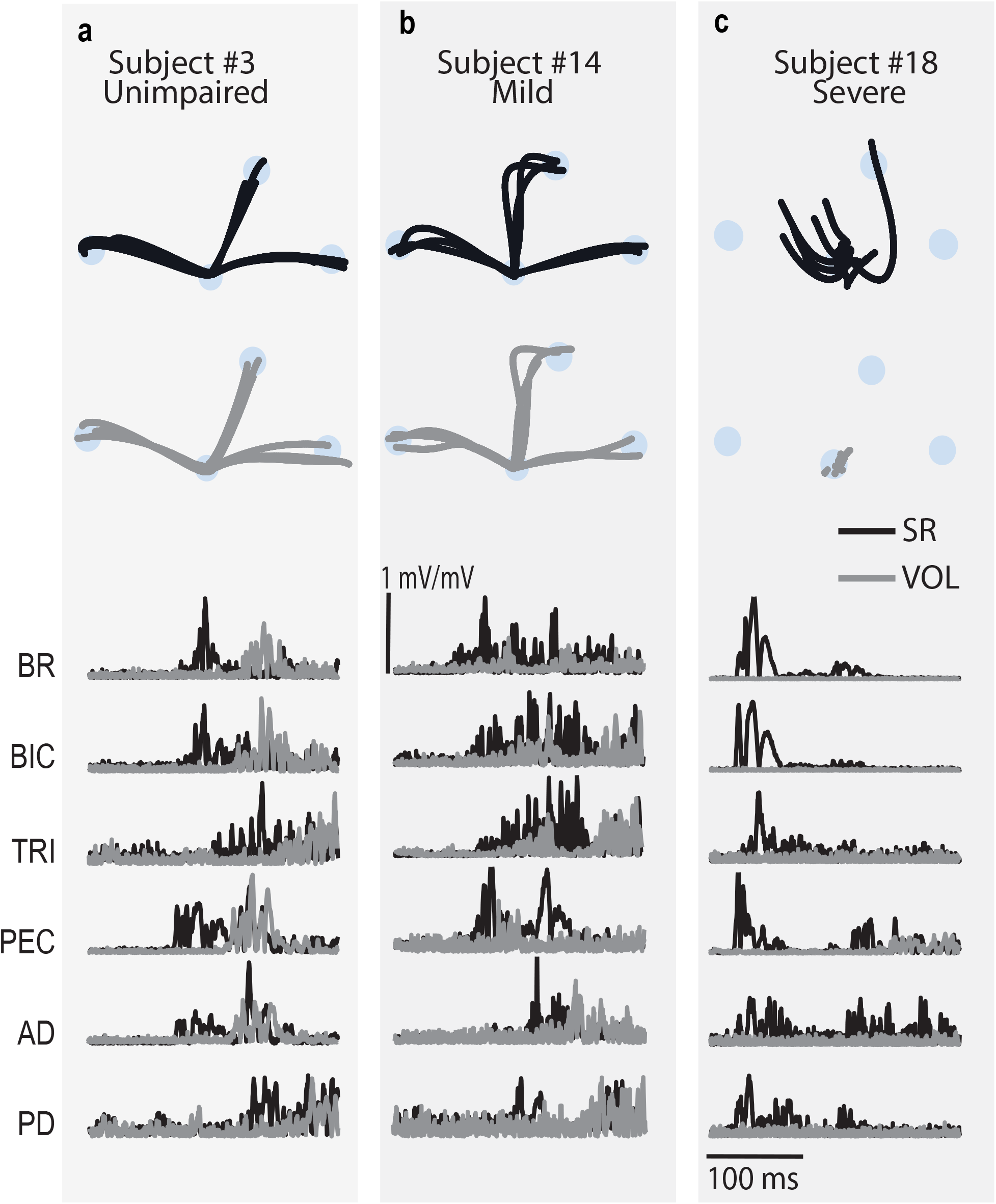
Representative data. VOL (gray) and SR (black) movement trajectories and EMG are depicted for individuals with unimpaired arm (**a**), mild (**b**) and severe (**c**) stroke.

Group results showed that SR trials had an increase in probability of muscle activity (severe/moderate & mild), larger muscle activity amplitude (all groups) and faster muscle activity onset (all groups) (Fig. 3). The probability of muscle activation in all muscles (BR, BIC, TRI, PEC, AD, PD) in severe/moderate group was increased in SR trials (*avg* Δ= 0.34 ± 0.057, all: P < 0.0001). Probability of muscle activity was also increased for some muscles in mild group during SR trials (*avg* Δ for BR, BIC and PEC = 0.12 ± 0.06, all: P < 0.05) but not for TRI, AD and PD (all: P > 0.057). As expected, the probability of activation during SR trials was not different from VOL in the unimpaired arm (all: P > 0.059), as indicated by the 96.83% average activation rate across all during the VOL condition. SR EMG amplitudes were larger for all populations (severe/moderate: *avg* Δ= 1.26 ± 0.34 *mV*/*mV*, mild: *avg* Δ= 0.21 ± 0.038 *mV*/*mV*, unimpaired: *avg* Δ= 0.22 ± 0.029 *mV*/*mV*, all: P ∼ 0). It is of note that during SR EMG amplitudes were greater than the maximum voluntary EMG activity in four muscles (BR, BIC, PEC, and AD) in the severe/moderate group. Typical of SR, SR muscle onset latencies were also faster than VOL for all populations (severe/moderate: *avg* Δ= 324.74 ± 13.33 *ms*, mild: *avg* Δ= 128.45 ± 11.21 *ms*, unimpaired: *avg* Δ= 99.55 ± 6.20 *ms*, all: P ∼ 0).

**Fig. 3.**
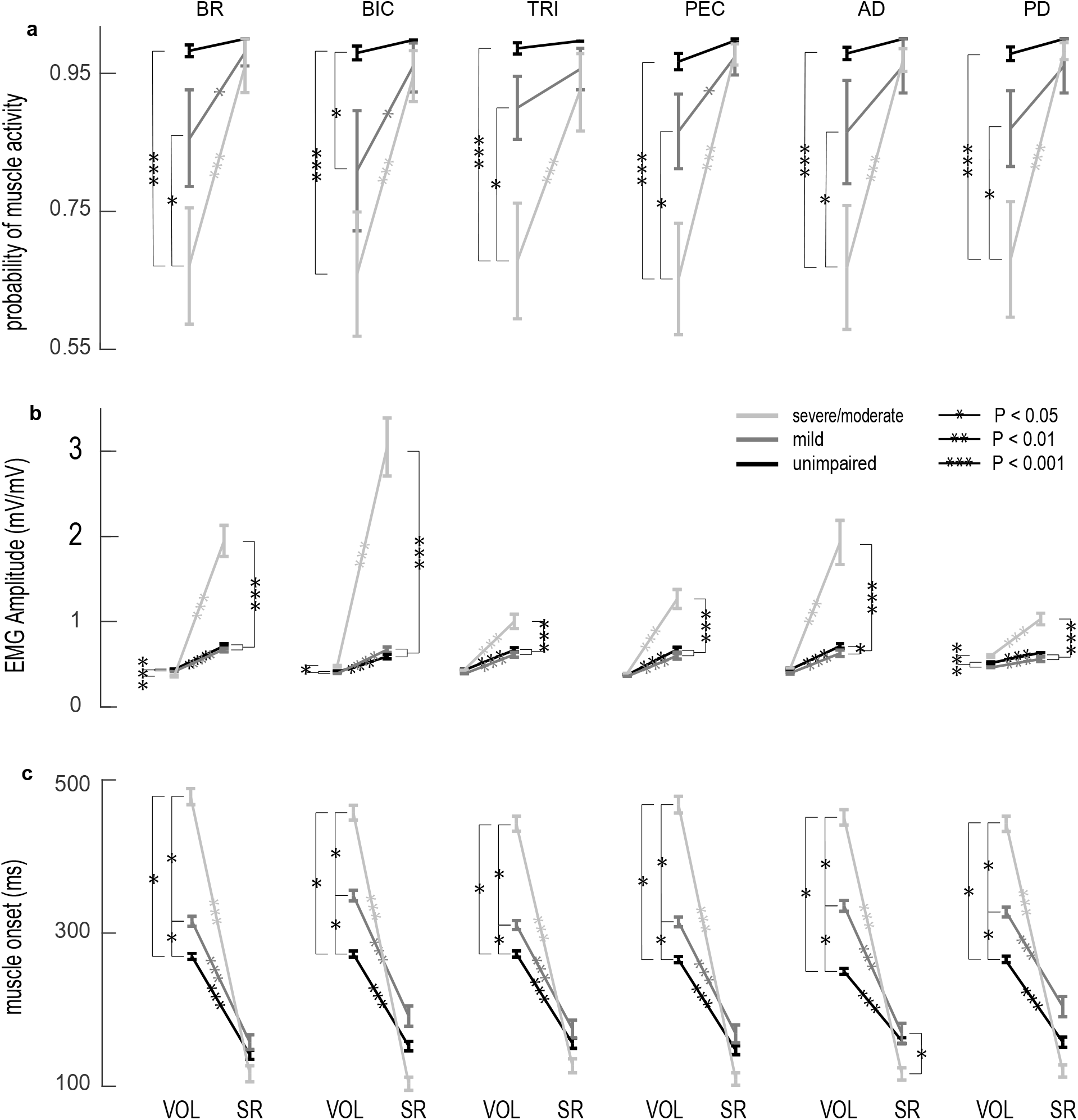
Group results of movement metrics. Movement onset (**a**), normalized distance traveled (**b**), final position error (**c**), and deviation from linearity (**d**) during VOL and SR are compared within each population: unimpaired, mild, and severe/moderate.

Despite changes in muscle activity, metrics of movement (distance, final error, linear deviation) were largely the same between SR and VOL trials, (Fig. 4) except in severe/moderate stroke who had larger reaching distances (Δ = 0.074 ± 0.0023 mm/mm, P = 0.001) and more deviation from linearity (Δ = 0.035 ± 0.019 *ms*, P = 0.048). Final error was not different between SR and VOL trials across all populations (all: P > 0.15) and deviation from linearity was also not different in unimpaired and mild groups (all: P >0.10). Movement distance was also not different for unimpaired (P = 0.11) and mild (P = 0.13). Consistent with SR, movement reaction time was faster for all populations (severe/moderate: Δ = 190.17 ± 9.34 *ms*, mild: Δ = 111.25 ± 6.33 *ms* and unimpaired: Δ = 78.23 ± 8.63 *ms*, all: P ∼ 0).

**Fig. 4.**
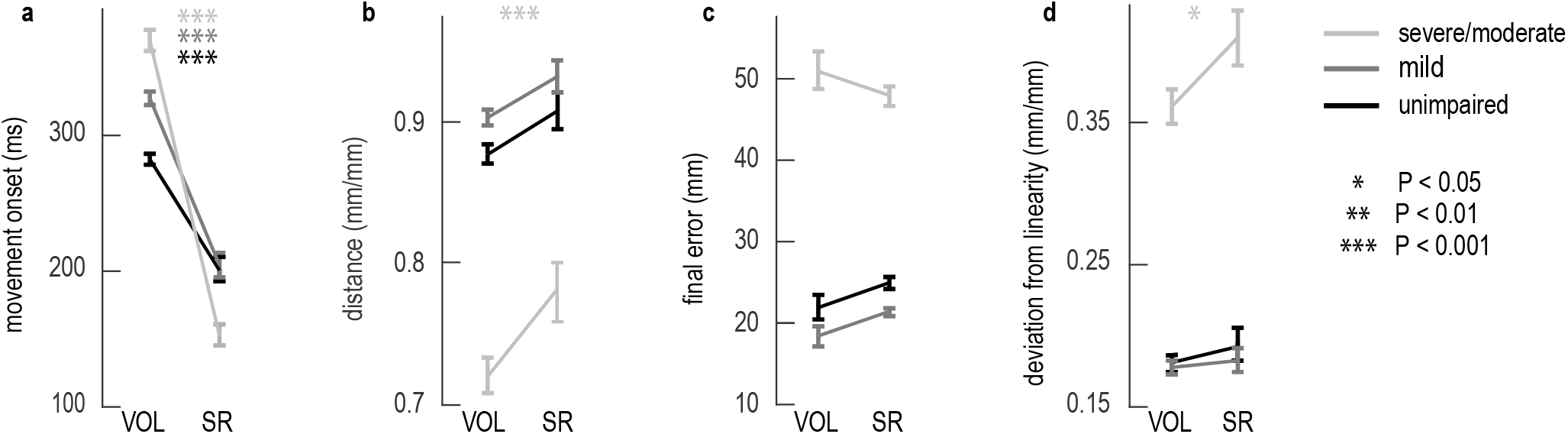
Group results of muscle activity metrics. The probability of muscle activity (**a**), EMG amplitude (**b**), and muscle activation onset (**c**) are compared across populations: unimpaired, mild, severe/moderate.

While VOL trials were different across the populations in terms of probability of muscle activity and onset latency, these metrics were largely the same across populations during SR trials. During VOL, the probability of muscle activity was smaller in the severe/moderate group compared to both mild (*avg* Δ = 0.20 ± 0.09, *all P* < 0.03, except for BIC P = 0.072) and unimpaired (*avg* Δ= 0.31 ± 0.09, P < 0.0001) but during SR trials the probability of muscle activity was not different across all of the groups (all: P > 0.2). Similarly, VOL muscle onset in the unimpaired group was faster than mild (*avg* Δ = 42.03 ± 6.37*ms*, P < 0.041) and faster than severe/moderate (*avg* Δ = 245.29 ± 10.19 *ms*, P < 0.025) but during SR trials there was no difference in muscle onset across all the groups (all: P > 0.11) except for AD which had a faster onset in the severe/moderate group (Δ = 38.02 ± 11.41 *ms*, p < 0.01). Finally, unlike the previous metrics, SR EMG amplitudes were larger in severe/moderate compared to mild (*avg* Δ = 1.00 ± 0.17, *all*: P < 0.001) and unimpaired (*avg* Δ= 1.03 ± 0.18, *all*: P < 0.001).

Classic startle resulted in no movement or muscle activity in the mild and unimpaired but 45% of the severe/moderate group showed movement – always directed towards the body. No movement or muscle activity was reported for classic startle in unimpaired arms or mild impaired arms (all traces from all subjects depicted, Fig. 5a and 5b); however, 5 out of 11 severe/moderate subjects moved out of the HOME circle during classic startle and generated muscle activity during 93% of trials. All classic startle movements were directed towards the body (Fig. 5c).

**Fig. 5.**
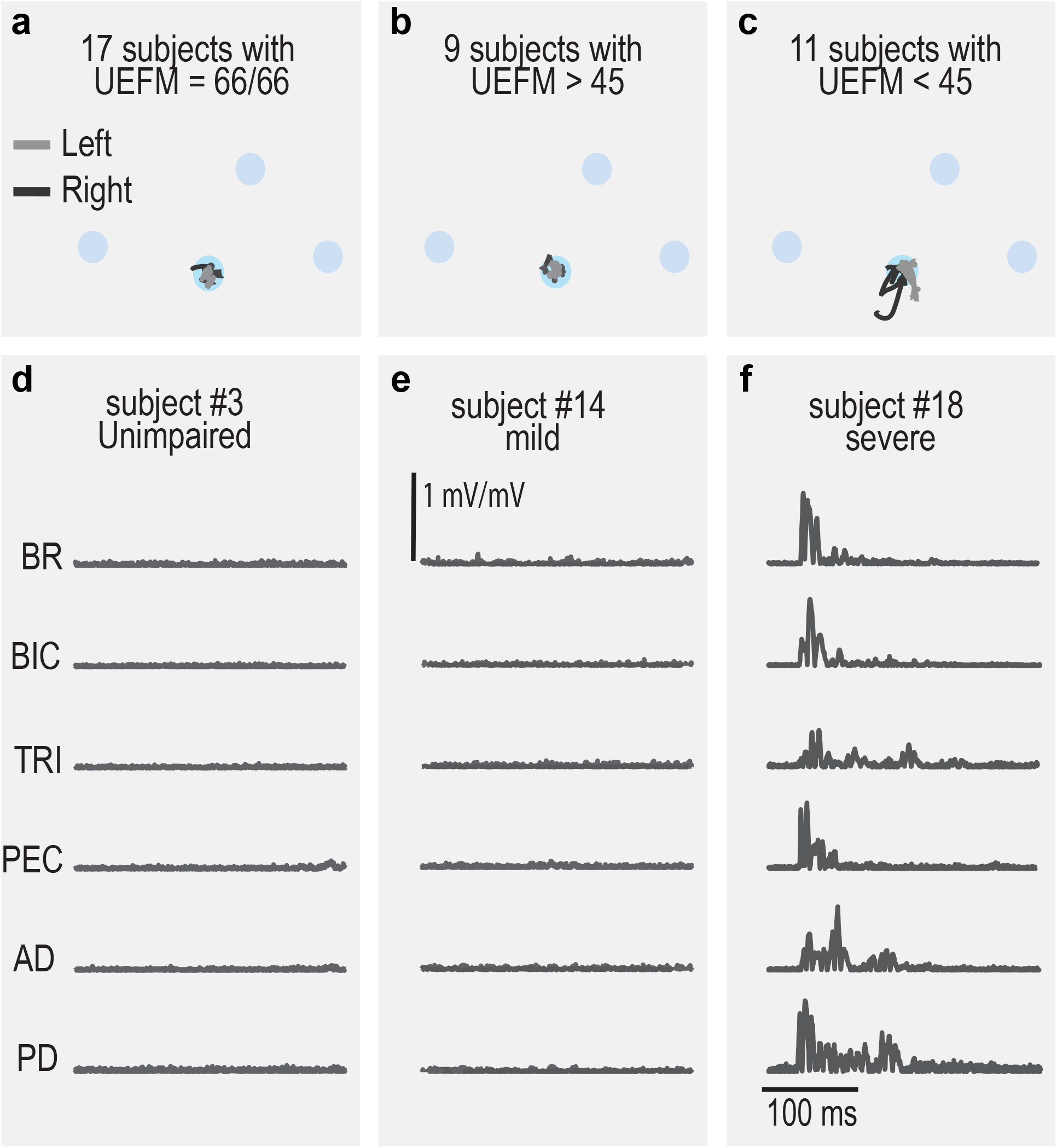
Classic startle data. All movement trajectories recorded during classic startle trials are depicted for unimpaired (**a**), mild (**b**), and severe/moderate (**c**) groups. In contrast, representative EMG from a single classic startle trials for each population is presented.

## Discussion

The objective of this study was to evaluate the impact of SR on unrestricted, two-dimensional reaching tasks in individuals with a wide range of impairment – in particular severe and moderate stroke. SR was robustly present in subjects across all impairment levels. Similar to prior reports in unimpaired adults (Ossanna et al. 2019) VOL and SR trials were nearly identical in terms of probability of muscle activity and movement metrics in the mild and unimpaired groups; however SR had a significant effect on muscle activity in the severe/moderate group. all metrics changed (larger muscle activity, decreased muscle latency), SR had the most profound effect on the probability of muscle activity. During VOL, the severe/moderate group activated their muscles during only 62.44% of trials compared to 97.85% during SR trials. This additional muscle activity allowed severe/moderate subjects to increase reaching distance but did not translate to a decrease in final error suggesting that the additional movement was not always directed towards the appropriate target. In conclusion, these results indicate that SR allowed the activation of paralyzed muscle in severe/moderate patients allowing further reaching but has little impact on mild patients during point-to-point reaching.

### SR and Rehabilitation

To our knowledge, SR has only been evaluated in 28 stroke survivors. This study nearly doubles the number of subjects evaluated to date to 51 and is the first SR study to include individuals with bilateral stroke. SR was robustly present in most (87%) of the stroke survivors in this study (Upper Extremity Fugl-Meyer’s ranging 8 - 66 and Modified Ashworth scores ranging: 0 - 4) and was well tolerated i.e. no adverse reactions were reported. It is of note that SR is also occasionally absent in young, neurological intact individuals (Kirkpatrick et al. 2018; Ossanna et al. 2019); thus it is uncertain if the absence of SR is related to the stroke or just a naturally occurring phenomenon.

The most profound impact of SR was the increase in probability of muscle activation, and the amplitude of that activation, in the severe/moderate stroke group. SR increased the probability of muscle activation 34% and increased activation amplitude to nearly two times the maximum voluntary capacity of the individual in 66% of muscles, suggesting SR is a profound way to activate paralyzed muscle. This impact was immediate – occurring in a single session and required no training. Furthermore, the increased muscle activity allowed severe/moderate individuals to generate larger reaching movements – though the final error was not altered indicating that these reaches were not necessarily functional. Still, this study evaluated a single SR session (∼1 hour) with only 45 SR trials per arm. A larger dose of SR over the course of several days may be required to see significant changes to target acquisition. In mild stroke survivors, SR increased the amplitude of muscle activity, but no other metrics of movement were altered. This indicates that either SR was not an appropriate target for mild stroke survivors or that SR did not generate quantitative improvements in tasks that were already readily performed by the subject.

Before exploring SR as a therapeutic target, it is important to assess the potential risk to severe patients. It has been noted in different studies that SR generates abnormal and inappropriate flexor activity that disrupts movement (Honeycutt and Perreault 2014; Tazoe and Perez 2017; Choudhury et al. 2019). Further, this activity increases with impairment. This raises concerns that more extended sessions of SR stimulation might induce neuroplastic changes that could further excite flexor synergies or spasticity. An alternative hypothesis is that SR might allow release spasticity through endogenous stimulation of paralyzed muscle. One of our most effective therapies to release spasticity is functional electrical stimulation (FES) of spastic muscle (Sabut et al. 2011). This works by releasing microfilament bridges and allowing the influx of ions to the muscle. SR might allow an endogenous stimulation of muscle which could potentially release muscle activity. Still, neither of these hypotheses has been explored and further evaluation is needed to determine whether SR is a safe and effective method to treat neurological impairment and spasticity following stroke.

### Abnormal Flexor activity

This work adds to the growing evidence that SR has a larger impact in flexor muscles, particularly in severe stroke. Previous studies have shown that SR extension movements are interrupted by abnormal flexor activity that arrives at the beginning of movement. This activity pulls the subject away from the intended target (Brown et al. 1991; Honeycutt and Perreault 2012) and increases with impairment level (Honeycutt and Perreault 2014; Honeycutt et al. 2015; Choudhury et al. 2019). Similarly, the abnormal activity in the flexors is larger in patients with severe spasticity (Choudhury et al. 2019). In this report, SR EMG amplitude was larger than the maximum voluntary EMG in flexor (BR, BIC, PEC, and AD) but not extensor (TRI, PD) muscles in the severe/moderate group. It is important to note that the flexor activity was not simply a result of the classic startle response. The classic startle response rarely activates extensor muscles (Honeycutt and Perreault 2012) and the classic startle trials collected here never resulted in movement directed towards the target. During SR trials extensor muscles demonstrated higher probability of muscle activity with larger response and most importantly, the movements directed towards the target. Taken together, these results indicate that SR in severe/moderate subjects was distinct from the classic startle response. However, the abnormally large flexor activity was present, and it disrupted movements in the severe/moderate population which raises concerns about the use of this technique in rehabilitation.

## Conclusion

The objective of this study was to evaluate the impact of SR on unrestricted, two-dimensional reaching tasks in individuals with a wide range of impairment – in particular, severe and moderate stroke. SR was robustly present in subjects across all impairment levels, but the most profound impact of SR was the increase in probability of muscle activation, and the amplitude of that activation, in the severe/moderate stroke group which led to increased reaching distance. This effect was immediate and required no training. Still enthusiasm is tempered because though SR increased reaching distance and muscle activity, the final error was not decreased which indicates that reaching movements were not necessarily functional. Future studies should evaluate the impact of longer-term SR exposure on both benefits of SR (increased probability of muscle activity) as well as any potential safety concerns (increased flexion synergies).

## Data Availability

all data is available in human mobility lab.

## Acknowledgements

This study was made possible through funding from the National Institutes of Health (R00 HD073240). Many thanks to members of the Human Mobility Lab for the guidance, assistance, and support throughout this research study.

